# Differences in epidemiology of enteropathogens in children pre- and post-rotavirus vaccine introduction in Kilifi, coastal Kenya

**DOI:** 10.1101/2022.04.28.22274420

**Authors:** Charles N. Agoti, Martin D. Curran, Nickson Murunga, Moses Ngari, Ester Muthumbi, Arnold Lambisia, Simon DW Frost, Barbara Blacklaws, D. James Nokes, Lydia N Drumright

## Abstract

**Background:** In July 2014, Kenya introduced the Rotarix® vaccine into its national immunization program. The impact of this vaccination programme on the local epidemiology of enteropathogens is unclear.

**Methods:** The TaqMan Array Card (TAC) was used for screening for 28 different enteropathogens in 718 stools from children less than 13 years of age who presented with diarrhea and were admitted to Kilifi County Hospital, coastal Kenya, in 2013 (before vaccine introduction) and in 2016-2018 (after vaccine introduction). The differences between pre- and post-Rotarix® vaccination periods were examined using univariate and multivariable logistic regressions.

**Results:** In 665 specimens (92.6%), one or more enteropathogens were detected, while in 323 specimens (48.6%), three or more enteropathogens were detected. There was a significant increase in the proportion of samples containing enteroaggregative *Escherichia coli* (35.7% vs 45.3%, *p=0.014*), cytomegalovirus (4.2% vs 9.9%, *p=0.008*), *Vibrio cholerae* (0.0% vs 2.3%, *p=0.019*), *Strongyloides species* (0.8% vs 3.6%, *p=0.048*) and *Dientamoeba fragilis* (2.1% vs 7.8%, *p=0.004*) post-vaccine introduction. Sapovirus detection decreased significantly (7.6% vs 4.0%, *p=0.030*) post-vaccine introduction. The proportion of samples that tested positive for rotavirus group A did not statistically differ between the pre- and post-vaccine periods (27.4% vs. 23.5%, *p=0.253*).

**Conclusions:** In this setting, the burden of childhood enteropathogen infection was high both pre- and post-rotavirus vaccination introduction, with some specific changes in the burden of enteropathogens in hospitalized children after rotavirus vaccination introduction.

## INTRODUCTION

In 2016, there were approximately 446,000 deaths worldwide caused by diarrheal illnesses among children <5 years, with the majority occurring in low-income countries [1, 2]. Rotavirus group A (RVA) was estimated to be responsible for approximately 128,000 of these deaths.

The World Health Organization (WHO) recommended inclusion of RVA vaccines in national immunization programs (NIPs) of all countries in 2009. Kenya included Rotarix® vaccination in its National Immunization Program [3] in July 2014, with doses at 6 and 10 weeks of age. Based on antigen testing, Kilifi County Hospital (KCH), Kenya, reported a 57% reduction in rotavirus hospitalizations in the first year after vaccine introduction and an 80% reduction in the second year among children <5 year-olds [4]. KCH found 64% vaccine effectiveness for children aged < 5years [5], which is similar to other low-income settings in sub-Saharan Africa [3, 4], but lower than high-income settings [5, 6].

Over 30 enteropathogens can cause diarrhea. In addition to a reduction in RVA-induced diarrhea, a referral hospital in Central Kenya observed a 31% decrease in all-cause diarrheal hospital admissions in the first year and a 58% decrease in the second year [7]. While RVA has clearly been responsible for a large burden of severe diarrheal cases among children <5 years-old, many other enteropathogens contribute to severe diarrheal illness in Kenya [8-11] but their epidemiology, both pre- and post-RVA vaccine introduction, is under-studied. Previously, we examined change in prevalence of 5 enteric viruses, including RVA, pre and post RVA vaccination demonstrating an increase in norovirus GII and a decrease in RVA, however this study did not examine all causes of diarrheal illness [12].

Through the use of a custom TaqMan Array Card (TAC), this study aimed to compare the epidemiological patterns of 28 different enteropathogens in children < 13 years-old admitted to KCH with diarrhea, pre- and post-RVA vaccine introduction.

## METHODS

### Study design and population

KCH is a referral facility that primarily serves residents of Kilifi County, Kenya [13]. From September 2009, a surveillance study of rotavirus was established at KCH pediatric ward targeting children <13 years old who presented with diarrhea (defined as passing three or more loose stools in a 24-hour period) [14]. A single stool specimen was collected within 48 hours of admission and immediately put into -4°C before transferring to the adjoining KEMRI-Wellcome Trust Laboratories for long-term storage at -80°C. The current retrospective analysis targeted samples collected in 2013 (pre-vaccine period) and 2016-2018 (post-vaccine period). During these periods, sample collection was interrupted 6 times by healthcare worker strikes; 3 times each pre- and post-vaccine introduction [15].

### Sample processing

Total nucleic acid was extracted from stool using two different approaches. The *cador* Pathogen 96 QIAcube HT Kit (QIAGEN, Hilden, Germany) with 200 µL liquid or 200 mg solid stool was used on samples from 2013. The QIAamp Fast DNA Stool Mini Kit (QIAGEN, Hilden, Germany) with 200 µL or 200 mg stool after bead-beating and centrifugation to remove debris was used on samples from 2016-2018. An extraction blank was included in every processing batch for quality control. The total nucleic acid extracts (NAE) were stored at -80°C for up to 130 weeks and defrosted once, only for the enteropathogen diagnostics.

### TaqMan Array Card (TAC) analysis

A custom TaqMan Array Card (TAC) spotted with lyophilized target-specific primers and probes was used to detect enteropathogens from the total NAE. The GASTRO v4.0K TAC was designed by MC at Public Health England (PHE), Cambridge, England [16] and manufactured by ThermoFisher, USA. GASTRO v4.0K screened for 28 enteropathogens: 12 viruses, 10 bacteria, 5 protozoa and 1 helminth (supplementary Table 1). All TAC targets were validated previously by PHE and include 16S bacterial and 18S RNA as internal controls, and bacteriophage MS2 as an external control. A Rotarix^®^ vaccine-specific assay targeting the NSP2 gene was included to identify RVA vaccine shedding due to prior vaccination. The molecular probes were labeled at the 5’ end with 6-carboxyfluorescein (FAM) reporter dye and NFQ-MGB quencher dye at the 3’ end [16].

For each TAC, 8 samples were tested (48-wells per sample). A mix of 60 µL of nuclease-free water, 25 µL of TaqMan Fast Virus 1-Step MasterMix (ThermoFisher, USA) containing ROX reference dye and 15 µL of NAE was prepared and loaded onto the TAC sample portal. TACs were spun twice at 1200 rpm for two minutes and sealed. TACs were processed using the Quantistudio-7-Flex Real-time PCR System (Thermofisher, USA). The thermocycling conditions were; 50°C for 5 minutes followed by 45 cycles of 95°C for 1 second and 60°C for 20 seconds. Extraction blanks and no template controls were included once in every 10 runs for quality control.

Amplification data were analysed by the cycle threshold (Ct) method (QuantiStudio(tm)Real-Time PCR software v.1.1). A threshold value of 0.2 florescence units was applied to all our analyses and baseline range set to automatic. The amplification curves for all presumptive positive samples were visually inspected prior to recording them as positive. Pathogen prevalence was evaluated at three different Ct cut-off levels (<40.0, <35.0 and <30.0).

### Clinical data

Data were collected using a standard clinical research form (CRF) entered into an electronic database [17]. The CRF included admission date, discharge date, presenting signs and symptoms, co-morbidities, and discharge diagnosis and outcome. Diarrhea was categorized as acute if the duration <7 days, prolonged if 7-13 days, persistent if 14-28 days, and chronic if >28 days. Diarrheal severity was assessed by Vesikari Clinical Severity Score (VCSS) [18]. All samples were clinically tested for rotavirus using a commercial ELISA antigen detection kit (ProsPeCT, Oxoid, Thermo Scientific. All pediatric admissions were offered HIV testing using two rapid antibody tests according to national guidelines.

### Analyses

We compared clinical and demographic characteristics of (i) included and excluded children and (ii) those recruited pre- and post-RVA vaccination using chi-squared analysis and Fisher’s exact test for categorical variables and Wilcoxon-rank sum and t-tests for continuous variables. As this was the first use of this TAC in Kenya, we descriptively explored different lower limits of detection (Ct ≤ 30, 35, 40) for a positive result. Using Ct ≤35 as the lower limit of detection, pathogen prevalence was calculated as the proportion positive over the total tested with 95% confidence intervals (CI) calculated using the exact method. We assessed differences in pathogen presence in pre-/post-RVA vaccination periods by univariate logistic regression. Any enteropathogen that was marginally different (P<0.1) in univariate analyses, as well as rotavirus (due to epidemiological interest), were included in multivariable logistic regression analyses adjusted for age, sex, Vesikari score, HIV-status, hospital length of stay (LoS), and fatality. Separate multivariate logistic regression analyses per pathogen were conducted due to the high prevalence of co-infections. The analyses were performed in Stata version 16.1 and R version 3.6.3.

### Ethical considerations

Before sample collection, informed written consent was gained from each child’s parent/guardian. The Scientific and Ethics Review Unit (SERU) at Kenya Medical Research Institute, Nairobi, approved the study protocol (Protocol #3624).

## RESULTS

### Participant characteristics

1,317 KCH pediatric admissions with diarrhea were eligible for inclusion during study periods (2013, 2016-2018). Of these, 1250 (95%) were unique patients; 718 (57.4%) provided an adequate stool sample for TAC processing. Children who did and did not have a stool sample were similar except those with a stool sample: (i) were more likely to have reported vomiting at admission (*p=0.001*); (ii) were hospitalized for longer (mean 5.8 days versus 4.3 days, *p <0.001*); (iii) were more likely to have been discharged alive (*p<0.001*); (iv) were more likely to have or have been exposed to HIV (p=0.049); and (v) were less likely to be included in 2017 and 2018 than in 2013 and 2016 (p<0.001) (**Table 1)**.

**Table 1.**
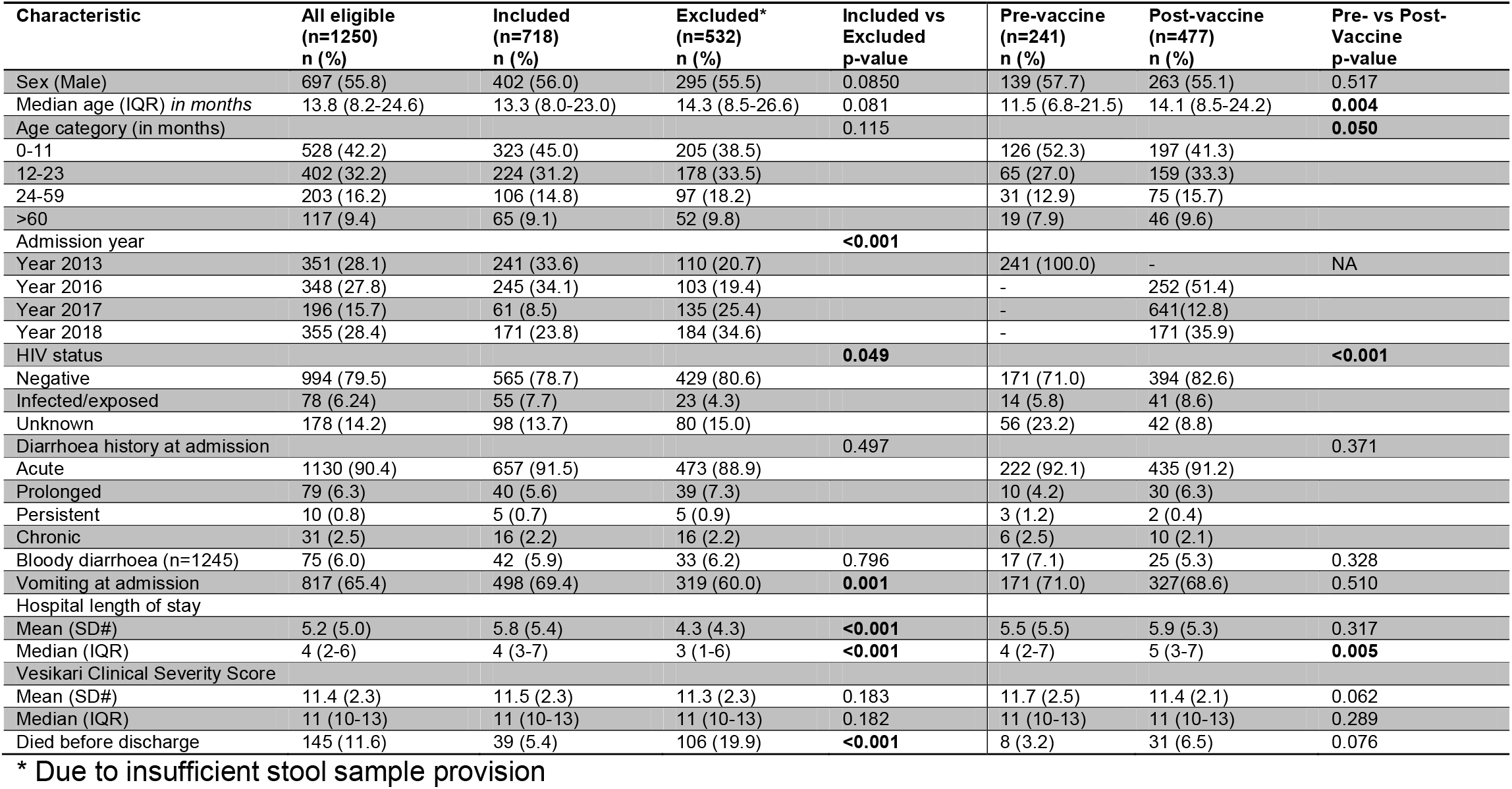
Demographic and clinical characteristics of the eligible, included and excluded participants and, among participants, pre-vaccine versus the post-vaccine introduction period comparisons.

| Characteristic | All eligible<br>(n=1250)<br>n (%) | Included<br>(n=718)<br>n (%) | Excluded*<br>(n=532)<br>n (%) | Included vs<br>Excluded<br>p-value | Pre-vaccine<br>(n=241)<br>n (%) | Post-vaccine<br>(n=477)<br>n (%) | Pre- vs Post-<br>Vaccine<br>p-value |
| --- | --- | --- | --- | --- | --- | --- | --- |
| Sex (Male) | 697 (55.8) | 402 (56.0) | 295 (55.5) | 0.0850 | 139 (57.7) | 263 (55.1) | 0.517 |
| Median age (IQR) *in months* | 13.8 (8.2-24.6) | 13.3 (8.0-23.0) | 14.3 (8.5-26.6) | 0.081 | 11.5 (6.8-21.5) | 14.1 (8.5-24.2) | **0.004** |
| Age category (in months) |  |  |  | 0.115 |  |  | **0.050** |
| 0-11 | 528 (42.2) | 323 (45.0) | 205 (38.5) |  | 126 (52.3) | 197 (41.3) |  |
| 12-23 | 402 (32.2) | 224 (31.2) | 178 (33.5) |  | 65 (27.0) | 159 (33.3) |  |
| 24-59 | 203 (16.2) | 106 (14.8) | 97 (18.2) |  | 31 (12.9) | 75 (15.7) |  |
| >60 | 117 (9.4) | 65 (9.1) | 52 (9.8) |  | 19 (7.9) | 46 (9.6) |  |
| Admission year |  |  |  | **<0.001** |  |  |  |
| Year 2013 | 351 (28.1) | 241 (33.6) | 110 (20.7) |  | 241 (100.0) | - | NA |
| Year 2016 | 348 (27.8) | 245 (34.1) | 103 (19.4) |  | - | 252 (51.4) |  |
| Year 2017 | 196 (15.7) | 61 (8.5) | 135 (25.4) |  | - | 641 (12.8) |  |
| Year 2018 | 355 (28.4) | 171 (23.8) | 184 (34.6) |  | - | 171 (35.9) |  |
| HIV status |  |  |  | **0.049** |  |  | **<0.001** |
| Negative | 994 (79.5) | 565 (78.7) | 429 (80.6) |  | 171 (71.0) | 394 (82.6) |  |
| Infected/exposed | 78 (6.24) | 55 (7.7) | 23 (4.3) |  | 14 (5.8) | 41 (8.6) |  |
| Unknown | 178 (14.2) | 98 (13.7) | 80 (15.0) |  | 56 (23.2) | 42 (8.8) |  |
| Diarrhoea history at admission |  |  |  | 0.497 |  |  | 0.371 |
| Acute | 1130 (90.4) | 657 (91.5) | 473 (88.9) |  | 222 (92.1) | 435 (91.2) |  |
| Prolonged | 79 (6.3) | 40 (5.6) | 39 (7.3) |  | 10 (4.2) | 30 (6.3) |  |
| Persistent | 10 (0.8) | 5 (0.7) | 5 (0.9) |  | 3 (1.2) | 2 (0.4) |  |
| Chronic | 31 (2.5) | 16 (2.2) | 16 (2.2) |  | 6 (2.5) | 10 (2.1) |  |
| Bloody diarrhoea (n=1245) | 75 (6.0) | 42 (5.9) | 33 (6.2) | 0.796 | 17 (7.1) | 25 (5.3) | 0.328 |
| Vomiting at admission | 817 (65.4) | 498 (69.4) | 319 (60.0) | **0.001** | 171 (71.0) | 327 (68.6) | 0.510 |
| Hospital length of stay |  |  |  |  |  |  |  |
| Mean (SD#) | 5.2 (5.0) | 5.8 (5.4) | 4.3 (4.3) | **<0.001** | 5.5 (5.5) | 5.9 (5.3) | 0.317 |
| Median (IQR) | 4 (2-6) | 4 (3-7) | 3 (1-6) | **<0.001** | 4 (2-7) | 5 (3-7) | **0.005** |
| Vesikari Clinical Severity Score |  |  |  |  |  |  |  |
| Mean (SD#) | 11.4 (2.3) | 11.5 (2.3) | 11.3 (2.3) | 0.183 | 11.7 (2.5) | 11.4 (2.1) | 0.062 |
| Median (IQR) | 11 (10-13) | 11 (10-13) | 11 (10-13) | 0.182 | 11 (10-13) | 11 (10-13) | 0.289 |
| Died before discharge | 145 (11.6) | 39 (5.4) | 106 (19.9) | **<0.001** | 8 (3.2) | 31 (6.5) | 0.076 |

\* Due to insufficient stool sample provision

The children who had provided sufficient stool had a median age of 13.3 months (IQR: 8.0-23.0). Clinical and demographic characteristics of children from pre-(2013) and post-vaccine (2016-2018) introduction periods (n=718) were similar except that those from the pre-vaccine period were (i) significantly younger (11.5 vs 14.1 months, p=0.004); (ii) less likely to have known HIV exposure, but more likely to have unknown HIV status (p<0.001); and (iii) had a lower median hospital stay (4 vs 5 days, p=0.005) than those from the post-vaccine period (Table 1).

### Pathogen prevalence

Of the 718 specimens analyzed by TAC, using a polymerase chain reaction (PCR) cycle threshold cut-off of ≤40.0 Ct (which was considered an appropriate cut-off based on clinical samples from the UK [16, 19]) one or more enteropathogen was detected in 94.0% of the children (n=675). The five most common pathogens at the ≤40.0 Ct cut off value were: enteroaggregative *Escherichia coli* (EAggEC) 44.6%; enterovirus 33.0%; enteropathogenic *Escherichia coli* (EPEC) 32.3%; RVA 25.1%; and parechovirus 20.8% **(Figure 1a)**. When the limit of detection was lowered to ≤30 Ct (high pathogen burden), 12/28 of detected pathogens decreased in prevalence by a factor of 2 or more (**Figure 1c)**. The largest declines were observed with verotoxogenic *E. coli* (VTEC) (11 fold), *Dientamoeba fragilis* (8.3 fold), cytomegalovirus (7.4 fold), *Strongyloides spp*. (5.0 fold), *Salmonella spp*. (4.0 fold), *Clostridium perfringens/difficile* (3.8 fold) and hepatitis E virus (3.0 fold). Only three pathogens, *D. fragilis*, hepatitis E virus, and VTEC, showed a reduction by factor 2 or more when comparing a Ct value of ≤40 versus ≤35 as the limit of detection. On average, the prevalence of the detected pathogens decreased by 1.3 fold when the Ct cut-off was lowered from 40.0 to 35.0 **(Figure 1b)**. Notably, even with the most stringent Ct cut-off (≤30), the prevalence of RVA was unaffected: 25.1% (21.9-28.4) at Ct ≤40 versus 22.3% (19.3-25.5) at Ct ≤30. No samples tested positive for *Vibrio parahaemolyticus* and *Entamoeba histolytica* at any Ct detection limit. All subsequent analyses have used the Ct cut-off of ≤35.0 for all pathogens. Detection above this threshold has previously been considered clinically insignificant or irreproducible in studies from low-income settings [20, 21].

**Figure 1.**
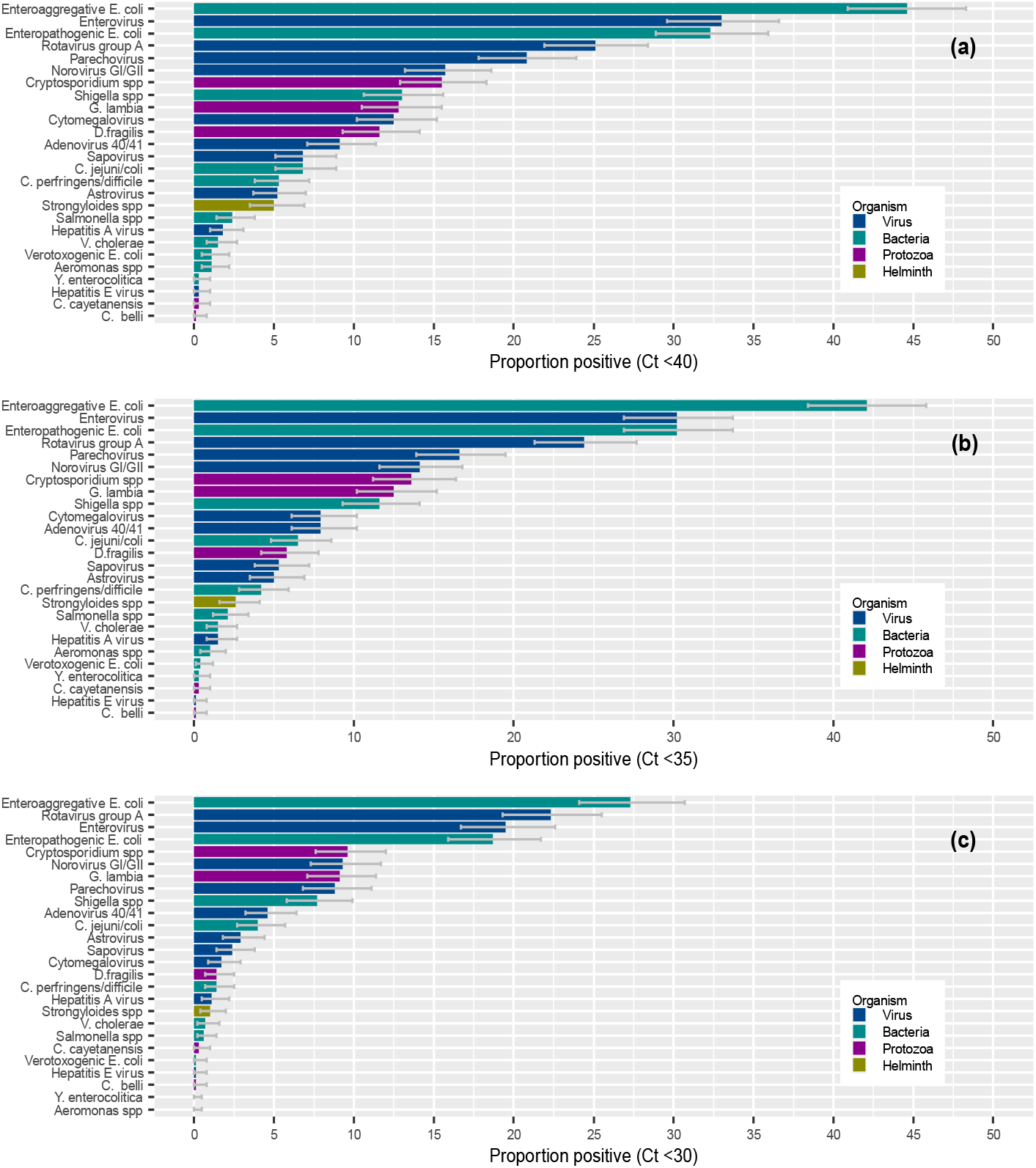
Prevalence of enteropathogens in stool samples from 718 children hospitalised at KCH at different PCR cycle threshold (Ct) cut-offs. The bars represent organism type and include 95% confidence interval errors bars for the proportions. Panel (a) prevalence of the detected enteropathogens when applying a Ct cut-off of ≤40 to define positives. Panel (b) prevalence of the detected enteropathogens when applying a Ct cut-off ≤35 to define positives. Panel (c) prevalence of detected the enteropathogens when applying a Ct cut-off ≤30 to define positives.

### Pathogens pre- and post-vaccine initiation

Among the 718 samples, 241 (33.6%) were from the pre-vaccine period (2013) and 477 (66.4%) were from the post-vaccine introduction period (2016-2018), (**Table 2**). In univariate logistic regression analyses, cytomegalovirus (4.2% vs 9.9%, OR=2.53, *p=0.010*), EAggEC (35.7% vs 45.3%, OR=1.49, *p=0.009*), *D. fragilis* (2.0% vs 7.8%, OR=3.97, *p=0.004*) and *Strongyloides spp*. (0.8% vs 3.6%, OR=4,42, *p=0.048)* all increased in detection from pre- to post-vaccine periods, whereas sapovirus decreased in detection (7.8% vs 4.0%, OR=0.49, *p=0.030*).

**Table 2.**
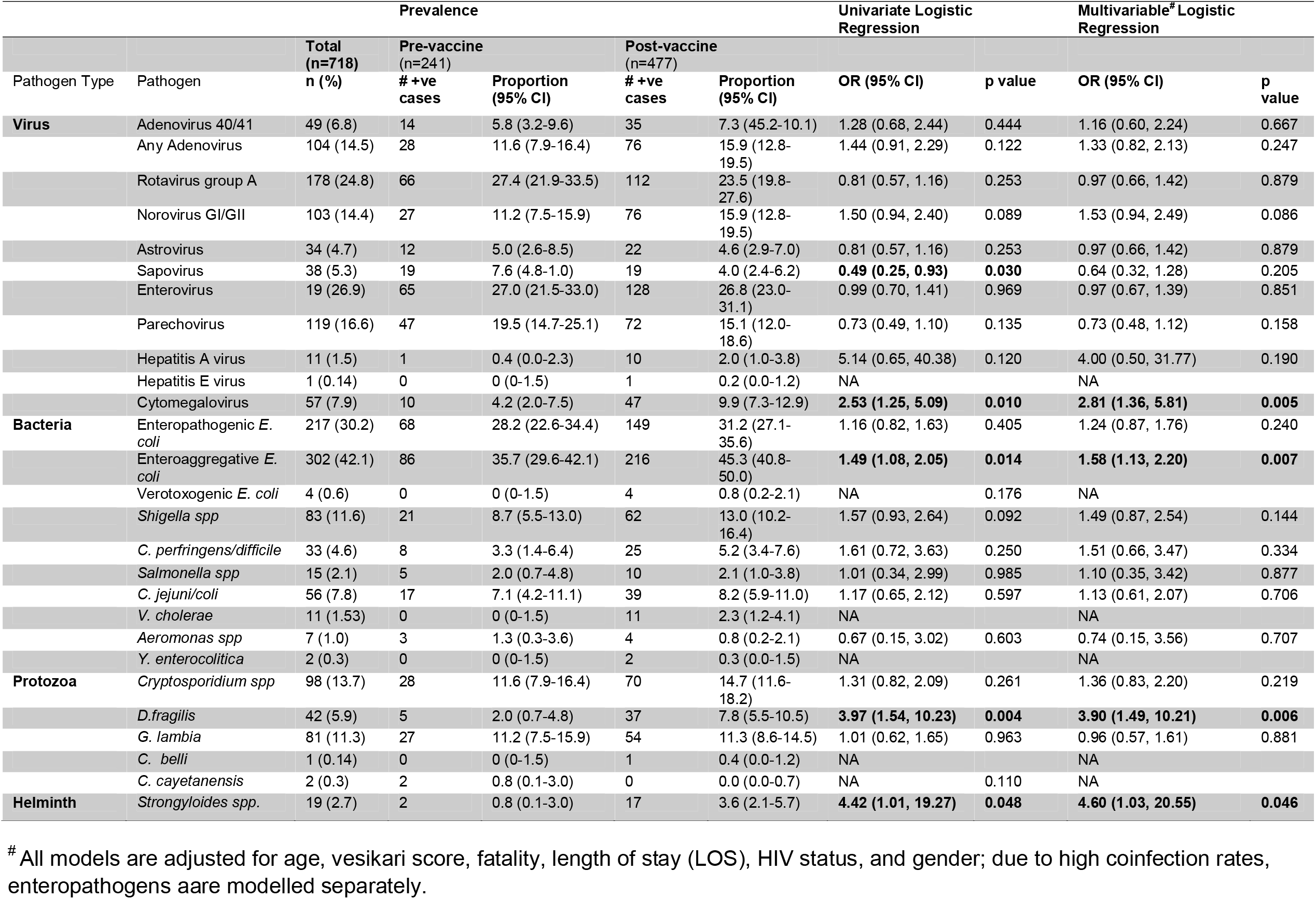
Univariate and multivariate logistic regression comparing enteropathogen infection in the pre- vs post-rotavirus vaccine introduction periods (n=718)

In multivariable models adjusting for age, Vesikari score, fatality, gender, LOS, and HIV-status, cytomegalovirus (OR=2.81, 95% CI 1.36, 5.81), EAggEC (OR=1.58, 95% CI 1.13, 2.20), *D. fragilis* (OR=3.90, 95% CI 1.49, 10.21), and *Strongyloides spp*. (OR=4.60, 95% CI 1.03, 20.55) were all detected among significantly more patients in the post-vaccine than in the pre-vaccine period. Sapovirus no longer demonstrated a significant difference in cases detected after adjusting for covariates **(Table 2)**.

No differences were observed in the prevalence of RVA in the hospitalized children in the pre-post-RVA vaccination time-periods. In sub-analyses of rotavirus infection, receipt of RVA vaccine was not associated with rotavirus infection in multiply adjusted models including age (data not shown). Both lack of reduction in RVA prevalence and lack of association with vaccine status were still consistently observed when restricting analyses to children >5 years old. No significant changes were observed for any of the other remaining pathogens.

### Co-infections

Only 53 children (7.4%) had no enteropathogens detected. Among the rest (n=665), 148 (22.3%) were mono-infected, while 323 (48.6%) had ≥3 pathogens detected. Including all children, 72% (n=517) had ≥2 enteropathogens detected. Five or more enteropathogens were detected in 84 children (11.7%). A greater number of different enteropathogens were detected per child in the post-vaccine compared to pre-vaccine period (mean: 2.7 vs 2.3 respectively, *p=0.0025*).

### Characteristics of rotavirus positive samples

The distribution of Ct values for RVA positive samples based on the TAC analysis is shown in Figure 2, in relation to (a) previous RVA ELISA test result, (b) sample period, (c) vaccination status, and (d) age group. We found that the median Ct values among older children are significantly lower than those among younger children, regardless of the sampling year.

**Figure 2.**
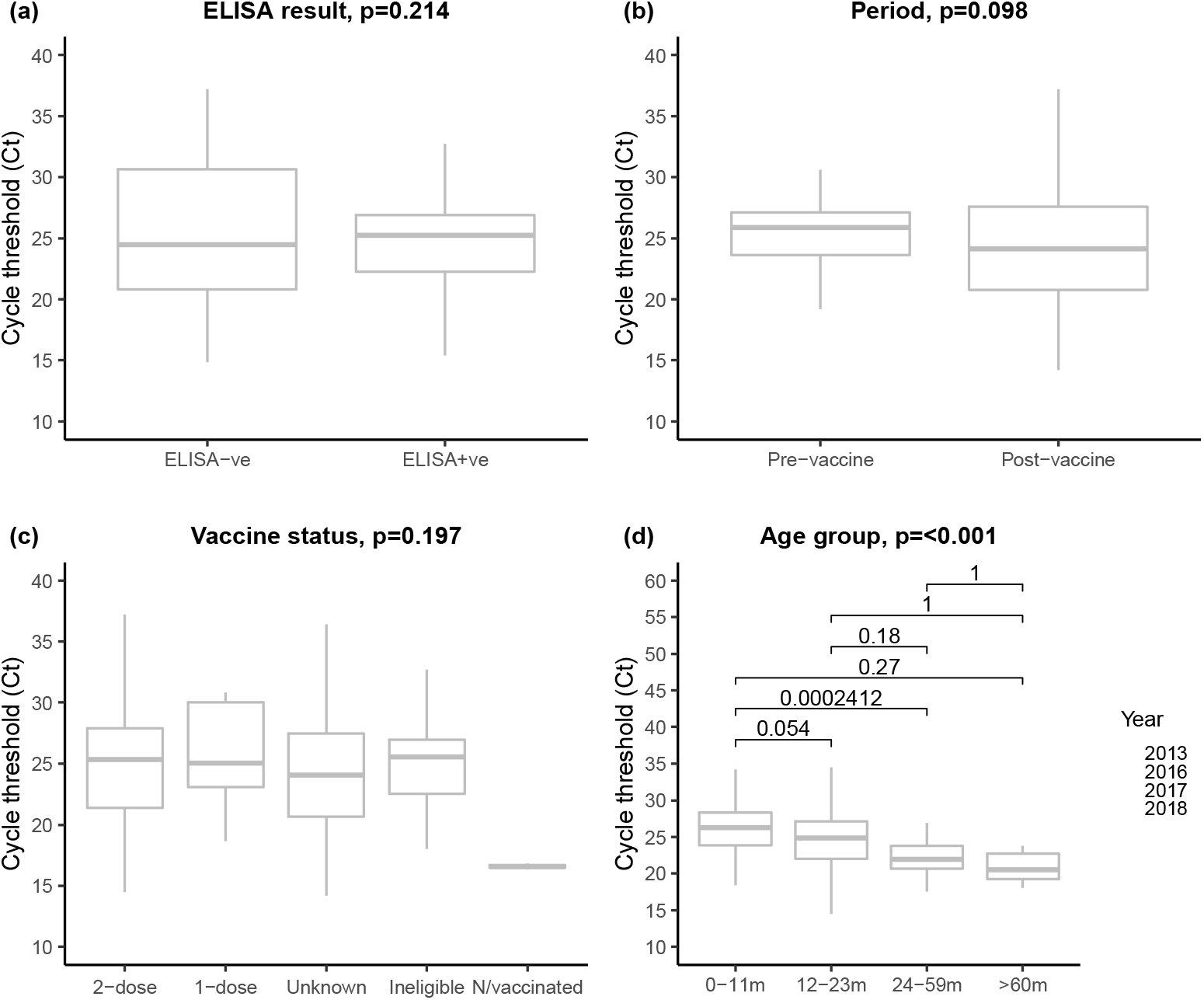
Boxplots examining the relationship between the observed RVA detection cycle threshold (Ct) from TAC assay and (a) previous enzyme-linked immunosorbent assay (ELISA) result on the same samples, (b) period of sample collection, (c) rotavirus vaccination status, and (d) age group in stool samples from 718 children at KCH. P-values are derived from an independent t-test, one-way ANOVA, and where applicable Kruskal Wallis was used.

## DISCUSSION

Our study shows very high prevalence of enteropathogens among hospitalized children with diarrhea, extensive co-infections, and differences in pathogen prevalence before and after rotavirus vaccine introduction. In addition, we observed no significant difference in RVA prevalence between pre- and post-vaccine introduction.

The Global Rotavirus Surveillance Network showed that RVA prevalence decreased in children under 5 years of age from 2008 and 2016 following national introduction of the RVA vaccine across 82 countries, although to a lesser extent among low-income countries [24]. Using controlled interrupted time series analysis, >50% decline in RVA cases were observed in two sites in Kenya following Rotarix® introduction [22]. One of these sites was KCH. In a previous analysis, using different viral detection assays, we also observed a reduction in RVA among children <5 years old [12]. Our study did not find any reduction in RVA prevalence among KCH children; however, there were some key differences between our study and the previous studies. First, to detect RVA infection, we used a TAC, which is considerably more sensitive than the immunoassay (EIA) used in other studies [23]. Secondly, our study included children up to 13 years of age, whereas the other studies included only children under 5. In the previous Kenyan studies, children were sampled between 2014 and 2017 [22] and 2003, 2013, 2016, and 2019 [12], which does not overlap entirely with our study. The inclusion of older children and the use of TAC provide evidence of a general prevalence of RVA in children, as opposed to only the vaccine eligible children [24].

The most frequently detected enteropathogens in our study in descending order were; EAggEC, EPEC, enterovirus, RVA, parechovirus, Norovirus, *Cryptosporidium spp*., *Shigella spp*. and *Giardia lambia*. In the Global Enteric Multicenter Study of children <5 years-old, the most common pathogens associated with diarrheal illness were RVA, enterotoxigenic *E. coli, Cryptosporidium spp*., *H. pylori*, norovirus GII, *Aeromonas spp*., adenovirus, EPEC, *Shigella spp*., *C. difficile*, and sapovirus [25]. Similarly, a study of bacterial diarrheal infections in <5 year-olds in Western Kenya revealed that *E. coli* and *Shigella* were the most common bacterial infections [26]. Our previous study only examining a handful of viral pathogens among children <5 years at KCH demonstrated low prevalence of viral infections, with RVA and adenovirus being the most common [12]. Although these studies took place in different geographic regions and/or used different methods of pathogen detection than ours, there is still consistency in the types of enteropathogens observed.

There have been a very few studies focusing on changes in multiple enteropathogen presentation after rotavirus vaccination [14, 22, 27-33]. We observed a significant increase in prevalence of cytomegalovirus, EAggEC, *D. fragilis* and *Strongyloides spp*. post-vaccination, as well as an increase in enteropathogen co-infection, suggesting that a reduction in RVA could result in a greater opportunity for other enteropathogens to fill the ecological niche previously occupied by RVA. Interestingly, a study among hospitalized children <5 years old in Tanzania examining the etiology of diarrheal illness post-rotavirus vaccine found RVA, ETEC, *Shigella, Cryptosporidium spp*. and astrovirus to be the five leading causes of diarrhea [34].

We demonstrated both a significant increase in prevalence of specific enteropathogens as well as co-infection following introduction of the rotavirus vaccine. While the detected enteropathogens could cause the diarrhea observed in these hospitalized children, it is unclear which are symptomatic illness versus long-term carriage. In order to understand changes in both burden and disease following RVA vaccine introduction, repeated measures studies on the same individuals including community controls are necessary. Such studies will also be important in understanding the longer-term impact of vaccination, carriage, and infection on children in LMIC settings.

This study had some limitations. Samples were collected more than two years prior to analysis, which could compromise sample integrity. This setting might have experienced a further change in enteropathogen burden since 2018 in part due to the COVID-19 pandemic. However, this does not invalidate the hypotheses generated by the findings regarding enteropathogen dynamics, but rather suggests that continuous surveillance is critical for adapting and maximizing diarrheal illness prevention. We did not analyze healthy controls to adjust for the background prevalence of the detected enteropathogens in this population. Of eligible children, 56.9% provided samples, which limited our ability to assess the entire population of hospitalised children with diarrhea. As a result of health worker industrial action in Kenya, enrollment was interrupted five times [15]. In the pre-vaccine and post-vaccine introduction years, the total nucleic acid extraction procedure varied, but we did not expect this to have significantly impacted enteropathogen prevalence. In spite of previous studies finding an association between pathogen quantities and diarrhea attribution, we did not attempt to model these associations in this study [21]. Finally, given the unbalanced time series nature of this study it is difficult to draw conclusions about enteropathogen dynamics following rotavirus vaccine introduction.

This study demonstrated increases in prevalence of EAEC, cytomegalovirus, *D. fragilis, Strongyloides spp*. and co-infections post-rotavirus vaccination, but little decrease in rotavirus infections in hospitalized children. Furthermore, we observed increased detection of RVA by TAC compared to ELISA. These findings highlight the need to use multiple detection arrays, such as TAC, for regular surveillance of hospitalized children and community controls in order to develop our understanding of differential observations of rotavirus vaccine efficacy in settings with different RVA prevalence in order to tailor the best vaccine approaches. Furthermore, such studies would provide insight into the clinical implications of the frequent co-infections among children in order to support high-quality interventions and individual patient care. Finally, our study suggests the need for broad diagnostic approaches with high sensitivity and specificity for enteropathogen surveillance and assessment of enteropathogen vaccination efficacy.

## Data Availability

The data described in this manuscript can be accessed by submitting a request form to our Data Governance Committee (http://kemri-wellcome.org/about-us/#ChildVerticalTab_15).

## Acknowledgement

We thank the study participants who provided the samples and members of the Virus Epidemiology and Control (VEC) group at KEMRI-Wellcome Trust Programme who collected the samples and performed the laboratory processing. We thank Prof. Philip Bejon for stimulating discussions about the work and comment provided on the initial draft manuscript. This study is published with permission from Director KEMRI.

