## Supplementray Table 1 for "Differences in epidemiology of enteropathogens in children pre- and post-rotavirus vaccine introduction in Kilifi, coastal Kenya"

**S1 Table.** Details of the targets on the Gastro v4.0K

| Pathogen Class | Pathogen/target | # of assays on Gastro v4.0K card | Gene (s) targeted | How positives were defined | Reference |
| --- | --- | --- | --- | --- | --- |
| Viruses | Adenovirus | 2 | Hexon gene | Both assays Ct<35.0 | [5] |
|  | Adenovirus 40/41 | 1 | Fiber gene | Ct <35.0 & Adenovirus +ve | [11] |
|  | Astrovirus | 2 | ORF2 & Capsid | Both assays Ct <35.0 | [5] |
|  | Sapovirus | 2 | RdRp | Either assay Ct <35.0 | [5] |
|  | Norovirus GI | 2 | ORF1/ORF2 | Either assay Ct <35.0 | [8] |
|  | Norovirus GII | 1 | ORF1/ORF2 | Ct <35.0 | [8] |
|  | Rotavirus group A | 2 | NSP3 | Both assays Ct <35.0 | [5] |
|  | Rotarix vaccine | 1 | NSP2 | Ct <35.0 and RVA +ve | [3] |
|  | Enterovirus | 2 | 5' UTR | Both assays Ct <35.0 | [2] |
|  | Hepatitis A virus | 1 | 5' UTR | Ct <35.0 | This study |
|  | Hepatitis E virus | 1 | ORF3 | Ct <35.0 | [9] |
|  | Parechovirus | 1 | 5' UTR | Ct <35.0 | This study |
|  | Cytomegalovirus | 1 | Immediate early gene | Ct <35.0 | This study |
| Bacteria | *Aeromonas hydrophilia* | 1 | aerolysin gene | Ct <35.0 | [6] |
|  | *Campylobacter coli/jejuni* | 1 | CadF gene | Ct <35.0 | [5] |
|  | *Campylobacter coli* | 1 | *ceuE gene* | Ct <35.0 & *Campylobacter spp* +ve | [1] |
|  | *Campylobacter jejuni#2* | 1 | mapA gene | Ct <35.0 & *Campylobacter spp* +ve | [1] |
|  | *Clostridium difficile* | 2 | GDH, ToxB | Both assays Ct <35.0 | [7] |
|  | *Clostridium perfringens* | 1 | α toxin | Ct <35.0 | 11 |
|  | *E. coli* EAEC | 1 | aggR gene | Ct <35.0 | This study |
|  | *E. coli* EPEC | 1 | *eae* | Ct <35.0 | [5] |
|  | *E. coli* VTEC | 2 | *vtx1* and *vtx2* | Either assays Ct <35.0 | [5] |
|  | *Salmonella spp* | 2 | *ttr* and *hilA* |  | 12 & 15 |
|  | *Shigella spp/*EIEC | 1 | *ipaH* |  | [5] |
|  | *Vibrio cholerae* | 1 | Tox R | Ct <35.0 | [5] |
|  | *Vibrio parahaemolyticus* | 1 | Tox R | Ct <35.0 | This study |
|  | *Yersinia enterocolitica* | 1 | lysP gene | Ct <35.0 | Liu et al 2 |
| Protozoa | *Cryptosporidium spp* | 2 | 18S rRNA and DNA J like gene | Ct <35.0 | [5] |
|  | *Cyclospora cayetanensis* | 1 | rRNA ITS2 | Ct <35.0 | [4] |
|  | *Cytoisospora belli* | 1 | 5.8S/ITS2 rRNA | Ct <35.0 | [10] |
|  | *Dientamoeba fragilis* | 1 | 5.8S rRNA | Ct <35.0 | 3 |
|  | *Entamoeba histolytica* | 1 | 18S rRNA | Ct <35.0 | [5] |
|  | *Giardia lambia* | 2 | 18S rRNA | Ct <35.0 | [5] |
| Helminth | *Strongyloides stercoralis* | 1 | 18S rRNA | Ct <35.0 | [12] |
| Controls | 16s Bacterial RNA | 1 | 16S rRNA | Ct <35.0 | This study |
|  | 18s Bacterial RNA | 1 | 18S rRNA | Ct <35.0 | Applied Biosystems |
|  | MS2 Bacteriophage | 1 | MS2g1 | Not applicable | [5, 8] |
| Total |  | 48 |  |  |  |

Ct stands for cycle threshold, ORF stands for open reading frame
